# Online media use and COVID-19 vaccination in real-world personal networks

**DOI:** 10.1101/2024.03.07.24303919

**Authors:** Iulian Oană, Marian-Gabriel Hâncean, Matjaž Perc, Jürgen Lerner, Bianca-Elena Pintoiu-Mihăilă, Marius Geantă, José Luis Molina, Isabela Tincă, Carolina Espina

## Abstract

**Background:** Most studies assessing the impact of online and social media usage on COVID-19 vaccine hesitancy predominantly rely on survey data, which often fails to capture the clustering of health opinions and behaviors within real-world networks. In contrast, research employing social network analysis aims to uncover the diverse communities and discourse themes related to vaccine support and hesitancy within social media platforms. Despite these advancements, there is a gap in the literature on how a person’s social circle, which combines online and offline interactions, affects vaccine acceptance.

**Objective:** We examined how online media consumption influences vaccination decisions within real-world social networks by analyzing unique quantitative network data collected from Romania, an Eastern European Union (EU) member state.

**Methods:** We conducted 83 face-to-face interviews with participants from a living lab in Leresti, a small rural community in Romania, employing a Personal Network Analysis (PNA) framework. This approach involved gathering data on both the respondents and individuals within their social circles (referred to as *social alters*). After excluding cases with missing data, our analysis proceeded with 61 complete personal networks. To examine the hierarchical structure of alters nested within ego networks, we utilized a mixed multilevel logistic regression model with random intercepts. The model aimed to predict vaccination status among alters, with the focal independent variable being the ego’s preferred source of health and prevention information. This variable was categorized into three types: traditional media, online media (including social media), and a combination of both, with traditional media serving as the reference category.

**Results:** In this study, we analyzed 61 personal networks, encompassing between 15 and 25 alters each, totaling 1280 alters with valid data across all variables of interest. Our primary findings indicate that alters within personal networks, whose respondents rely solely on online media for health information, exhibit lower vaccination rates (odds ratio [OR] 0.37, 95% CI 0.15-0.92; *P*=.03). Conversely, the transition from exclusive traditional media use to a combination of both traditional and online media does not significantly impact vaccination odds (OR 0.75, 95% CI 0.32-1.78; *P*=.52). Additionally, our analysis reveals that alters in personal networks with vaccinated egos are more likely to be vaccinated themselves (OR 3.75, 95% CI 1.79-7.85; *P*<.001).

**Conclusion:** Real-world networks combine offline and online human interactions with consequences on health opinions and behaviors. As individuals’ vaccination status is influenced by how their social alters use online media and vaccination behavior, further insights are needed to create tailored communication campaigns and interventions regarding vaccination in areas with low levels of digital health literacy and vaccination rates, as Romania exposes.

## Introduction

Currently, Coronavirus disease 2019 (COVID-19) is not as prominent on the public agenda, despite the fact that infections with the severe acute respiratory syndrome coronavirus 2 (SARS-CoV-2) are still occurring. At the 76th World Health Assembly, held between 21-30 May 2023, in Geneva, World Health Organization’s (WHO) chief Tedros Adhanom Ghebreyesus stated that the threat of other pandemics, either related to mutations of SARSLCoVL2 or other pathogens, is still present [1]. Moreover, all the debates regarding vaccination during the COVID-19 pandemic affected how people approach other vaccines, such as routine childhood vaccines [2], and raised questions about the impact of HPV and HBV vaccination [3]. Vaccine hesitancy has been one of the significant threats to public health, especially in developed nations [4]. The Strategic Advisory Group of Experts on Immunization (SAGE) of the WHO defined *vaccination hesitancy* as the *“delay in acceptance or refusal of vaccination despite the availability of vaccination services”* [5]. The SAGE working group adds that vaccination hesitancy is context-specific, a complex of beliefs and behaviors conditional on culture, history, and vaccine type.

While not a new phenomenon [6,7], the landscape in which vaccine hesitancy exists today is new. With the advent of the internet, social media, and people interconnected in a global network, which further scales up the phenomenon’s complexity, the WHO has stressed the importance of managing *infodemics* for efficient interventions against the COVID-19 pandemic [8]. The WHO defines an *infodemic* as “too much information, including false or misleading information in digital and physical environments during a disease outbreak” [9]. Working by misinformation or disinformation can lead to “confusion and risk-taking behaviors that can harm health. It also leads to mistrust in health authorities and undermines the public health response” [9]. Compared to traditional, legacy media (official news channels, radio, or newspapers), online media, in general, and social media, in particular, facilitate the spread of unverified medical information [10,11] and the creation of echo chambers where false information, conspiracy theories, and fears are reinforced [12].

Evidence on how online and social media usage affect COVID-19 vaccination is mixed. For instance, extant research indicates that these media types are either positively [13–19] or negatively correlated with hesitancy [20,21]. However, most studies presented in systematic reviews [22,23] or systematic reviews of reviews [24] suggest that online and social media usage tends to increase vaccination hesitancy. Similar conclusions were also drawn from analyses of cross-national surveys [13,19]. For studies that found a correlation between social media usage and vaccination acceptance, it is generally observed that such results typically emerge from specific samples, such as adolescents [25], who are more inclined to seek information online, or within unique cultural contexts like China [21], where the interaction with social media content significantly differs [26].

Supplementary results are also brought by scholars who analyzed the effects of social media in comparison with traditional media. Some studies have found that the usage of legacy media has a positive effect on vaccine acceptance, alongside the negative effect associated with social media [14,15,18]. In other studies, the focus was primarily on the correlation with traditional media, finding that the use of social media had no significant effect. [27,28].

Interestingly, one study found that not even traditional media as a whole category can correlate with vaccine acceptance. Viswanath et al. [20] distinguished between the usage of mainstream broadcast media and mainstream print media and found that only the latter positively affected the acceptance of the COVID-19 vaccine, the former being non- significant. In other research, no direct association was found between online or social media usage and vaccine hesitancy, or it was observed that the effects of social media vary due to indirect influences. Specifically, it was demonstrated that social media consumption can positively impact vaccine hesitancy when mediated by confidence in the vaccine’s safety. [27]. Lee and You also emphasize the importance of indirect associations [29]. Their study found that even if social media, in general, has a positive impact on vaccine hesitancy, this effect can be amplified by respondents’ perceived risk of vaccine-induced side effects or reversed through respondents’ perceived risk of getting infected with SARS-CoV-2.

Given that the majority of data on vaccination hesitancy during the pandemic was collected through standard surveys, statistical analyses have been conducted to assess the effects of media usage on vaccine hesitancy. These analyses control for various sociodemographic traits (such as gender, income, education, and ethnic group) and other individual-level characteristics, including trust in government, health experts, and vaccine confidence.

Nevertheless, it is crucial to recognize that information dissemination is not a one-way process from media outlets to isolated individuals. People’s opinions and attitudes are significantly shaped by their social circles, including family, peers, close friends, and other social groups they trust and interact with frequently. Opinions and behaviors regarding COVID-19 vaccination tend to be clustered within social groups rather than distributed randomly, as evidenced by studies [30,31]. This observation underscores the necessity of considering real-world social clusters when examining the impact of media usage on vaccination hesitancy. In line with this, the SAGE Working Group on Vaccine Hesitancy has acknowledged the significant role of social influence within the matrix of determinants for vaccine hesitancy [5].

Complementing studies that rely solely on standard survey data and focus on the attributes of individuals, network science introduces a more comprehensive approach by integrating attributes with relationships between units of analysis. Within this field, research on social actors falls into Social Network Analysis (SNA) and Personal Network Analysis (PNA), both of which conceptualize social actors as nodes and their relationships as ties. SNA concentrates on specific relationships within a defined population of nodes (e.g., students within a school or users reposting a particular hashtag) in a bounded context, adopting a *sociocentric* design that encapsulates all nodes within a singular network. In contrast, PNA explores the wider social circles of individuals through an *egocentric* design, focusing on the networks that revolve around central nodes (or *egos*) and their connections to various social *alters*. These alters may not be part of the same group boundaries, leading to multiple, distinct networks that often cannot be aggregated into a single network [32].

Studies employing Social Network Analysis (SNA) to examine COVID-19 vaccine hesitancy predominantly utilize datasets that capture user interactions within social media platforms.

These studies typically concentrate on analyzing the content of discussions related to vaccine hesitancy and identifying key participants or groups within these conversations through community detection algorithms. This approach allows for a nuanced understanding of the discourse dynamics and the social structures influencing vaccine hesitancy among online communities. Scholars found that while positive and negative discourses surrounding COVID-19 vaccination are present on social media platforms [26,33–36], they depend on political partisanship and the quality of sources [12,35]. Such studies are of great value for content analysis and mapping different communities inside online spaces. They identify online behavior patterns that, in the end, may affect day-to-day health decisions and bring insights into how to create better communication campaigns for vaccine acceptance.

Furthermore, they bring nuance in discussing the relationship between the usage of online platforms and vaccine hesitancy. Exposure to different sources in these online spaces (whom people choose to follow), the tendency to comment on posts presenting similar views [37], as well as prior biases (e.g., general level of distrust [28], trust in government [29], or political partisanship [20]), stress the idea that health outcomes are contextual and contingent on how people engage with these online spaces, and on what they bring to these spaces from offline influences.

However, research remains limited on the impact of real-world networks on COVID-19 vaccine hesitancy or acceptance [31] as well as on the extent to which these networks reflect online stances towards COVID-19 vaccination. This gap is significant given the potential for network structures and compositions to influence the transmission dynamics of the SARS-CoV-2 virus [38–40]. Past studies have demonstrated that health outcomes are associated with *assortative mixing*. In many instances, assortative mixing–wherein nodes with similar traits form connections more frequently–serves as an indication of a network’s structure and composition, emphasizing the tendency for similar individuals to be more closely inter- connected. Drivers of assortative mixing can be represented by *social influence* (or *contagion*), where behaviors or traits spread through the network; *social selection* (*homophily*), where individuals form connections based on similar characteristics; or *social context* (*confounding*), where external factors related to the environment or setting influence network formation [41,42]. Social contagion seeks to explain how nodal characteristics (behaviors, opinions, and other traits) change as a function of a node’s relations inside the network [43]. Homophily principles state that nodes tend to create ties with others who are similar to them [44]. Contextual influences refer to effects brought by macro-level changes (or lack of) in the social environment that influence individual or group behavior [41]. As examples of health outcomes related to the mechanisms mentioned above, we can name obesity [45] or the adoption of weight loss behaviors [46], the adoption of clean cooking methods [47], and how physicians adopt treatment plans and screening practices [48,49].

Regarding vaccination, previous studies have shown that network assortativity has a positive correlation with influenza vaccinations [50], opinions about COVID-19 vaccination [31], and that network structures can facilitate, in general, one’s acceptance or hesitancy through the influence of neighboring nodes [51].

In order to investigate how consumption of online media influences the vaccination decisions within real-world social networks, we use unique network data collected from Romania, a European Union (EU) member state situated in the Eastern Europe. As a general context, Romania has the second lowest COVID-19 vaccination rate when compared to other EU countries. The latest data recorded by Our World in Data on the 10th of June 2022 showed 41.27% of Romania’s population was fully vaccinated [52]. Such low vaccination rates are also troubling in the context of low digital literacy rates. Data from Eurostat show that in 2023, Romania was in last place in the EU when looking at the proportion of persons with basic or above basic overall digital skills–31.09% compared with the EU average of 60.55% for individuals who used the internet in the last 3 months [53]. Among EU states, in 2023, Romania was fourth in internet usage for social media platforms–84.35% compared with the EU average of 64.74% for individuals who used the internet in the last 3 months [54].

Internet usage for seeking health information is low but rapidly increasing. In 2022, Romania occupied the last position in this EU ranking–33.82% compared with the EU average of 57.74% for individuals who used the internet in the last 3 months [54]. In 2023, Romania was placed third to last in the EU ranking of seeking health information online–51.71% compared with the EU average of 61.51% for individuals who used the internet in the last 3 months [54].

Proxy indicators on Romanians’ digital health literacy show worrying facts about online media use. In 2023, the proportion of persons who checked the truthfulness of information from internet news sites or social media was the second lowest in the EU–11.24% compared with the EU average of 26.38% for individuals who used the internet in the last 3 months [55]. The proportion of persons who checked the truthfulness of such information by checking the sources or comparing it with other information found on the internet further reinforces the ranking of Romania as second lowest in the EU–8.96% compared with the EU average of 23.29% for individuals who used the internet in the last 3 months [55]. Eurostat figures about “evaluating data, information, and digital content” also indirectly show Romanians’ ability to identify misinformation or disinformation. The percentage of individuals reporting having seen untrue or doubtful information on the internet is the lowest in Romanian–32.83% compared with the EU average of 53.67% for individuals who used the internet in the last 3 months [55]. Such low numbers beg the question of whether Romanians truly do not encounter untrue information when using online media or cannot discern them from what is presented as accurate information.

Our study focuses on the critical role of online media in influencing vaccination behaviors, using PNA on real-world social data. It specifically examines how the exclusive use of online media for health information correlates with the vaccination tendencies among individuals’ social contacts in rural Romania (Eastern Europe). The investigation uncovers that within the personal networks of those who rely solely on online sources, there is a discernible decrease in vaccination rates. This finding shed light on the dynamics prevalent in regions marked by low COVID-19 vaccination uptake, limited digital health literacy, and overall digital literacy challenges.

Our work stands out for its application of unique quantitative data, offering a fresh perspective on the interaction between digital information consumption and health behavior, particularly in the context of vaccine hesitancy. Hopefully, we make a significant contribution by mixing online media usage and personal network influences on vaccination decisions, thereby filling a crucial gap in existing literature.

## Methods

### Data collection

The data presented in this study were collected during the pilot phase of the 4P-CAN project (Personalized cancer primary prevention research through citizen participation and digitally enable social innovation; HORIZON-MISS-2022-CANCER-01, project ID 101104432, program HORIZON) [56]. We collected data from one of 4P-CAN’s living labs in Leresti, a rural locality in Arges county, Romania (N=4557).

The data collection process took place between the 13th and 30th September 2023. We interviewed 83 persons following a Personal Network Analysis (PNA) design [32]. Study participants were recruited using a respondent-driven link-tracing sampling methodology [57,58]. At the time of the interview, every interviewee was at least 18 years old. A named institutional/licensing committee approved the research protocol. The Center for Innovation in Medicine (InoMed) Ethics Committee reviewed and approved all the study procedures (EC-INOMED Decision No. D001/19-01-2024). Before the interviews, we obtained informed consent from all study participants, ensuring their anonymity and alters’ anonymity.

Each participant responded to a questionnaire that collected information about their opinions, behaviors, and characteristics of individuals from their social circle. Each interviewee was asked to nominate up to 25 people with whom they interact frequently (face-to-face or via other communication methods) and who are at least 18 years old. The nominated persons could be *family members*, *partners, close friends, casual friends*, or *acquaintances*.

Afterward, the respondents were asked about certain characteristics of the people they nominated in their social circle. All interviews were conducted face-to-face, having an average duration of 80 minutes. The questionnaires were applied using Network Canvas [59], a PNA research software. Network analysis, in general, distinguishes between two data types: attribute and relational. Attribute data refer to characteristics measured individually for each person (*node*) within the network, whereas relational data describe propertied of the relations between the nodes, also known as *ties* [60]. In the context of PNA research, the nodes are the egos and alters, and ties refer to ego-alter and alter-alter relations. For each study participant (*ego*), we collected attribute data about them (e.g., age, marital status, various health behaviors, and opinions), attribute data about persons in their social circle (*alters*), relational data capturing ego-alter relationships, and relational data about alter-alter ties. It is important to note that all aforementioned data types were captured from the perspective of the egos.

### Attribute data

Of the attribute data, for both egos and alters, we collected information on their *sex* (1=female; 0=male), *age in years*, *being single* (1=yes; 0=no), and *COVID-19 vaccination status* (0=unvaccinated, 1=vaccinated). The education of egos and alters was measured by an ordinal scale capturing the last education level, with the values: 1=*no school*; 2=*less than primary school*; 3=*primary school*; 4=*less than secondary school*; 5=*secondary school*; 6=*arts & crafts school*; 7=*10 obligatory years*; 8=*high school - unfinished*; 9=*high school - finished (with diploma)*; 10=*post-high school (non-tertiary)*; 11=*bachelor’s degree or equivalent level*; 12=*master’s degree or equivalent level*; 13=*PhD or equivalent level*.

*Employment status* (1=employed; 0=other) was measured only for egos.

We asked the egos to provide details about their information sources related to medical topics. The egos had to respond to this question: *What are your sources of information about health and prevention (lifestyle)*? The respondents could select multiple answers from a) *central TV stations*; b) *local press*; c) *online, using Google, Bing, or other search engines*; d) *online, using social media (Facebook, TikTok, Instagram, etc.)*; e) *online influencers*. In our analyses, this was introduced as a factor variable distinguishing between those who use *only traditional media* (options *a, b,* or both), those who use *only online media* (*c*, *d, e,* or any combination of the three), and those who use *both types of media*. Reliance on traditional media only was used as a reference level. While we acknowledge a difference between reliance on social media for news and information and other internet mediums, we chose to group the egos in the general category of *online media* because only one respondent stated that they use only social media for information about health and prevention. Others who mentioned social media also mentioned search engines (and grouped as *online media* users) or legacy media (and grouped as *both media* users).

### Network data

Regarding ego-alter relations, we took into account the *emotional closeness* between egos and alters (*How emotionally close do you feel to this person?* with answer options: *not at all close*, *not very close*, *close*, and *very close*), and their *direct interaction frequency* (*How often do you typically meet with this person?* with answer options: *less than once a year*, *once a year*, *a few times a year*, *once a month*, *every two weeks, weekly, daily*). In both cases, the variables were binarized. For emotional closeness, 1 represents situations where the ego said they have a *very close* relation with the alter (0=other). For direct interaction frequency, 1 represents a situation where egos and alters meet at least twice a month (0=other). We used the product of the two binary variables to derive another binary variable, dubbed *ego-alter intensity*, where 1 represents the situation in which the ego feels very close to the alter, *and* they meet at least twice a month.

For the relations between alters, the participants were presented with the following question for each alter dyad: *Please tell me if these persons, even if they are related to each other, are acquittances, casual friends, or close friends.* From the alter-alter tie data, we derived *structural* network level and node level measurements after the ego was excluded from the personal network, its presence being redundant. All alter-alter ties were binarized, 1 representing the existence of a relation, while 0 denotes situations where alters do not know each other.

Node level structural measures are represented by alters’ *betweenness centrality* and *vaccination assortativity score*. Betweenness centrality quantifies the number of times a node acts as a bridge along the shortest path between two other nodes [61]. Thus, betweenness centrality can also be viewed as the power to control the flow of information and opinions about vaccination inside the personal network [31]. The betweenness scores were normalized to account for differing network sizes.

The *vaccination assortativity score* combines attribute and relational data to account for sub- groupings inside the personal network. It informs whether alters of similar traits (vaccinated or not) tend to share a social tie. This score was computed for each alter *i*, from the personal network of an ego *j*, taking the difference between the proportion of all of *j*’s alters that are connected to *i* and vaccinated, and the proportion of all of *j*’s alters that are vaccinated. In computing the proportions, *i* was not included. For example, let us suppose that ego *j* has nominated 25 alters, of whom 14 are vaccinated against COVID-19 and 11 are not. In this personal network, alter *i,* who is vaccinated, is directly connected with 9 other alters, of which 6 are vaccinated, and 3 are unvaccinated. The assortativity score for alter *i* (which is excluded from the computation) is: 6/9 - 13/24 = 0.666 - 0.542 = 0.142. This indicates that the alter’s *i* network neighbors have an above-average vaccination rate compared to the overall proportion of vaccinated alters in the personal network. Given that an alter’s vaccination status is measured via a binary variable (1=vaccinated; 0=unvaccinated), a positive correlation coefficient indicates assortative mixing (i.e., the alter’s vaccination status is related to the vaccination status of its direct neighbors in the network). The computation method was previously advanced to detect assortative mixing of opinions related to COVID- 19 vaccination [31] and processed food intake [62].

Network level measurements describe personal networks as a whole, accounting for the distribution of alter-alter ties. Network *size* represents the number of nodes inside a network (in our case, alters). *Density* sums up the proportion of present ties out of the theoretical number of ties (i.e., if all nodes are connected). Density can also be interpreted as a measure of cohesion inside the network [63]. Finally, the *number of components* is an indicator of network fragmentation. An unfragmented network is a network with one component where all nodes are directly or indirectly connected to each other. If a network has at least *n ≥ 2* components, it indicates the number of *n* clusters of alters that are separated from each other. Also, isolated nodes represent stand-alone components [61].

### Statistical analysis

Regarding the statistical modeling, the objective of our analyses was to assess the impact of egos’ online information consumption in predicting alters’ vaccination status: vaccinated or not. Given the nature of the dependent variable (binary) and the nested structure of the data– alters embedded in egos’ networks–a mixed multilevel logistic regression model with random intercepts [64]. In this framework, alter level data and ego-alter relations can be regarded as level one predictors (individual), while egos’ attributes and network level data are level two (group) [65]. This type of nesting must be controlled for, as the data about egos and alters are reported by egos for which unobserved covariates could influence the results [31]. Level one predictors include alters’ sex, education level, age, relationship status (being single), betweenness centrality, relation intensity with ego, and vaccination assortativity. Level two predictors include egos’ sex, education, age, employment status, and media use alongside network size, density, and number of strong components. To avoid scaling problems in the estimation of models, numeric variables were mean-centered and standardized. Even if education was measured via an ordinal scale, it was introduced in the statistical model as a numeric variable, given its large interval (from 1 to 13), and to let it have variation.

### Data exclusion

The initial sample was comprised of 83 respondents with varying network sizes and missing data about alters’ being vaccinated against COVID-19. To ensure that we have enough data at each level two group, we chose to focus our analyses only on those personal networks which have at least 15 alters for which data about vaccination was present and who are not isolates. Not being an isolate (having at least one tie with another alter) is an important condition, given that isolated alters, for which the assortativity score could not be computed (as they have no network neighbors), were dropped from the analysis. After filtering the data according to these conditions, we ended up with a subset of 64 personal networks. Out of the 64, the regression models and some bivariate analyses presented in Multimedia Appendix 1 took into consideration only 61 egos because data about media usage for health and prevention was missing for three of them. Egos’ relationship status was not introduced in the multilevel regression models, given the vast overrepresentation of those who are not single.

Out of the 61 egos, only 7 reported that they were single.

## Results

Table 1 presents descriptive statistics for characteristics of egos, personal networks, and alters. The majority of respondents (egos) are vaccinated (50/64, 78%). They have an average age of ≈ 53 years (SD 51.86), are almost equally split with respect to sex (33/64 are women), a large majority (56/64, 88%) are in a relation (being married or as a couple), and a little over half are employed (36/64, 56%). The average value for education is 9.7 (SD 1.78), in between secondary (finished high school) and post-secondary (non-tertiary) education on the ordinal scale. Distribution of responses for the type of media used for information about health and prevention shows that most egos use a combination of online and traditional media (29/64, 45%), 21 out of 64 use online media exclusively, and 11 use only traditional media as their information sources. The respondents’ social contacts (alters) have a similar average age of ≈ 53 years (SD 16.06), similar average education (mean 9.35, SD 1.93), and also approximately equally split on their sex (820/1561 are women). Also similar to egos, the vast majority of alters (1242/1561, 80%) are in a relation. Regarding their vaccination status, 64% of egos’ social contacts (991/1561) were reported as being vaccinated against COVID-19. For relations between egos and their social contacts, we report that for 22% (mean 0.22, SD 0.42) of ego-alter ties (347/1561), the respondents mentioned that they are ‘very close’ and meet at least ‘twice a month’ with the respective alter.

**Table 1.** Descriptive statistics for egos and alters.

| Characteristic | Values for ego & network data (n = 64) | Values for alter data (n = 1561) |
| --- | --- | --- |
| Age in years, mean (SD) | 53.33 (15.86) | 52.64 (16.06) |
| **Education level** |  |  |
| mean (SD) (NA) | 9.70 (1.78) (0) | 9.35 (1.93) (52) |
| median (IQR) (NA) | 10 (2) (0) | 9 (2) (52) |
| **Vaccinated against COVID-19, n (%)** |  |  |
| No | 14 (22) | 394 (25.24) |
| Yes | 50 (78) | 991 (63.48) |
| Missing | 0 (0) | 176 (11.27) |
| **Sex, n (%)** |  |  |
| Male | 31 (48) | 741 (47.47) |
| Female | 33 (52) | 820 (52.53) |
| **Being single, n (%)** |  |  |
| No | 56 (88) | 1242 (79.56) |
| Yes | 8 (12) | 318 (20.37) |
| Missing | 0 (0) | 1 (0.06) |
| **Being employed, n (%)** |  |  |
| No | 28 (44) | - |
| Yes | 36 (56) | - |
| **Media used for health and prevention, n (%)** |  |  |
| Traditional media | 11 (17) | - |
| Online media | 21 (33) | - |
| Both media | 29 (45) | - |
| Missing | 3 (5) | - |
| Ego-Alter intensity, mean (SD) | - | 0.22 (0.42) |
| Alter betweenness centrality, mean (SD) | - | 0.04 (0.07) |
| Vaccination assortativity, mean (SD) (NA) | - | 0.02 (0.11) (181) |
| Network size, mean (SD) | 24.39 (1.70) | - |
| Network density, mean (SD) | 0.65 (0.21) | - |
| Network components, mean (SD) | 1.16 (0.51) | - |

Summary statistics for personal networks’ structural features show that, on average, they are composed of 24 alters (56/64 have at least 24 alters, while the rest have between 15 and 23), with an average density of 0.65 (SD 0.21) and 1.16 number of strong components (SD 0.51), indicating high cohesiveness (density) and low fragmentation (components). A mean betweenness score of 0.04 (SD 0.07) (scaled by the network’s size) tells us that alters have a low level of control over the potential information flow between otherwise unconnected alters. The mean assortativity score for vaccination is 0.02 (SD 0.11), indicating that, on average, the proportion of an alter’s direct network neighbors who are vaccinated is similar to the overall proportion inside the personal network (minus the alter of reference).

More detailed descriptive statistics regarding the distributions of presented variables are in tables S3, S4, S7, and S13 provided in the Multimedia Appendix 1. In the supplementary file, we also report bivariate tests for some of the presented variables. First, looking at alters’ vaccination status contingent on egos’ vaccination status, we observe that the frequency distributions are not independent. Vaccinated respondents tend to have in their personal networks more vaccinated social contacts (*χ*^2^_1_ = 76.195; *P*<.001; table S15 in Multimedia Appendix 1 [MA1]). Second, the frequency distribution of alters’ vaccination status by egos’ type of media used for health information also indicates a non-independent distribution (*χ*^2^_2_ = 28.255; *P*<.001; table S16 in MA1). Egos using online media only present the highest proportion of unvaccinated alters (172/457, 37.6%), compared with egos using only traditional media (74/284, 29.8%) or both (140/616, 22.7%). However, it must be mentioned that there is no association between egos’ vaccination status and their preferred media for information about health and prevention distribution (*χ*^2^ = 2.542, Fisher’s *P*=0.315; table S17 in MA1). Two-sided t-tests of independent samples, comparing the average assortativity score grouped by alters’ or egos’ vaccination status, show that in both cases, assortative mixing is lower for unvaccinated alters (*t*_1378_=− 2.3835; *P*=0.02; table S19 in MA1) and unvaccinated egos (*t*_1378_=−3.5391; *P*<.001; table S21 in MA1). Similarly, by performing a chi-square test comparing the distribution of alters’ assortativity quartiles by egos’ media use and vaccination status, we found that the distribution is non-independent (*χ*^2^_15_ = 83.229; *P*<.001; table S22 in MA1). Egos who are unvaccinated and use only online media present the highest proportion of alters in quartiles 1 (33/106, 31.1%) and 2 (43/106, 40.6%).

In Table 2, we report the results of the full mixed multilevel logistic regression model, using alters’ vaccination status as an outcome variable for 61 groups and 1280 alters (i.e., cases without missing data on the variables included in the model). Egos’ relationship status (being single or not) was not included in the models due to its lack of variation. The intraclass correlation coefficient (ICC) of 0.2 indicates that we have within-group homogeneity and inter-group heterogeneity. The grouping dependence is also confirmed by the marginal and conditional *R*^2^ coefficients. The fixed effects explain 21.5% of the variance, while the random effects account for an extra 15.9% of the total variance. Regarding alters’ attributes, education increases the probability of an alter being vaccinated (odds ratio [OR] 1.87, 95% CI 1.58-2.22; *P*<.001), while being single lowers it (OR 0.67, 95% CI 0.46-0.96; *P*=.03).

**Table 2.** Mixed multilevel logistic regression predicting alters’ vaccination status (n=1280 alters and 61 egos)

| Predictors<sup>a</sup> |  |  | Full model<sup>b</sup> |  |  |
| --- | --- | --- | --- | --- | --- |
|  |  |  | OR<sup>c</sup> | 95% CI | *P* value |
| Intercept |  |  | 2.29 | 0.85-6.15 | 0.10 |
| Alter level (1) |  |  |  |  |  |
|  | Sex |  |  |  |  |
|  |  | Male | Reference |  |  |
|  |  | Female | 1.13 | 0.84-1.53 | 0.41 |
|  | Education |  | 1.87 | 1.58-2.22 | <0.001 |
|  | Being single |  |  |  |  |
|  |  | No | Reference |  |  |
|  |  | Yes | 0.67 | 0.46-0.96 | 0.03 |
|  | Alter age |  | 0.98 | 0.83-1.16 | 0.83 |
|  | Ego-Alter intensity |  | 0.92 | 0.62-1.35 | 0.66 |
|  | Alter betweenness |  | 1.03 | 0.88-1.20 | 0.71 |
|  | Vaccination assortativity |  | 1.17 | 1.01-1.35 | 0.04 |
| Ego level (2) |  |  |  |  |  |
|  | Sex |  |  |  |  |
|  |  | Male | Reference |  |  |
|  |  | Female | 1.04 | 0.57-1.91 | 0.89 |
|  | Education |  | 1.24 | 0.87-1.75 | 0.24 |
|  | Age |  | 1.02 | 0.73-1.42 | 0.92 |
|  | Being employed |  |  |  |  |
|  |  | No | Reference |  |  |
|  |  | Yes | 0.63 | 0.30-1.34 | 0.23 |
|  | Being vaccinated |  |  |  |  |
|  |  | No | Reference |  |  |
|  |  | Yes | 3.75 | 1.79-7.85 | <0.001 |
|  | Media type |  |  |  |  |
|  |  | Traditional | Reference |  |  |
|  |  | Online | 0.37 | 0.15-0.92 | 0.03 |
|  |  | Both | 0.75 | 0.32-1.78 | 0.52 |
| Network level (2) |  |  |  |  |  |
|  | Network size |  | 0.96 | 0.74-1.26 | 0.79 |
|  | Network density |  | 1.23 | 0.87-1.75 | 0.24 |
|  | Network components (strong) |  | 1.04 | 0.77-1.41 | 0.80 |

<sup>a</sup>With the exception of categorical variables and ego-alter intensity, all variables were mean-centered and scaled. The outcome variable remained in its original units.
<sup>b</sup>Model indices: Variance (SD)=0.83 (0.913); ICC=0.2; Observations=1280; Groups=61; Marginal $R^2$ =0.215; Conditional $R^2$ =0.374.
<sup>c</sup>Odds ratios.

Among egos’ attributes as level two predictors, we observe that being in the personal network of an interviewee who was vaccinated increases an alter’s chance of also being vaccinated (OR 3.75, 95% CI 1.79-7.85; *P*<.001). Network level structural characteristics (size, density, and number of components) have no significant effects on the outcome variable. Between the node level structural features (alter’s betweenness centrality and vaccination assortativity), only the variable accounting for assortative mixing presents a positive effect (OR 1.17, 95% CI 1.01-1.35; *P*=.04). For every standard deviation increase in the assortativity score, alters are 17% more likely to be vaccinated.

Egos’ media use for health and prevention information is our variable of interest, using traditional media as the reference category. Results indicate a negative effect for switching from the exclusive use of legacy media to the exclusive use of online media (OR 0.37, 95% CI 0.15-0.92; *P*=.03). The change from “traditional only” to both types of media has no effect (*P*=.52). For comparison, Figure 1 presents a visual summary of multiple models, with odds ratios and 95% confidence intervals. Model 2 represents a model where we include only predictors referring to egos and alters’ attributes. Model 3 is a model where we included only predictors that take into account relational data, such as ego-alter relation intensity, node-level structural features (betweenness centrality and vaccination assortativity), and network-level structural features. Table S23 from the Multimedia Appendix 1 presents in detail the full model (Model 4) in comparison with the null model (Model 1), and Models 2 and 3. In all models where included, the variable of interest–ego’s media use for health information– remained significant and with similar odds levels while controlling for the other level one and two factors.

**Figure 1.**
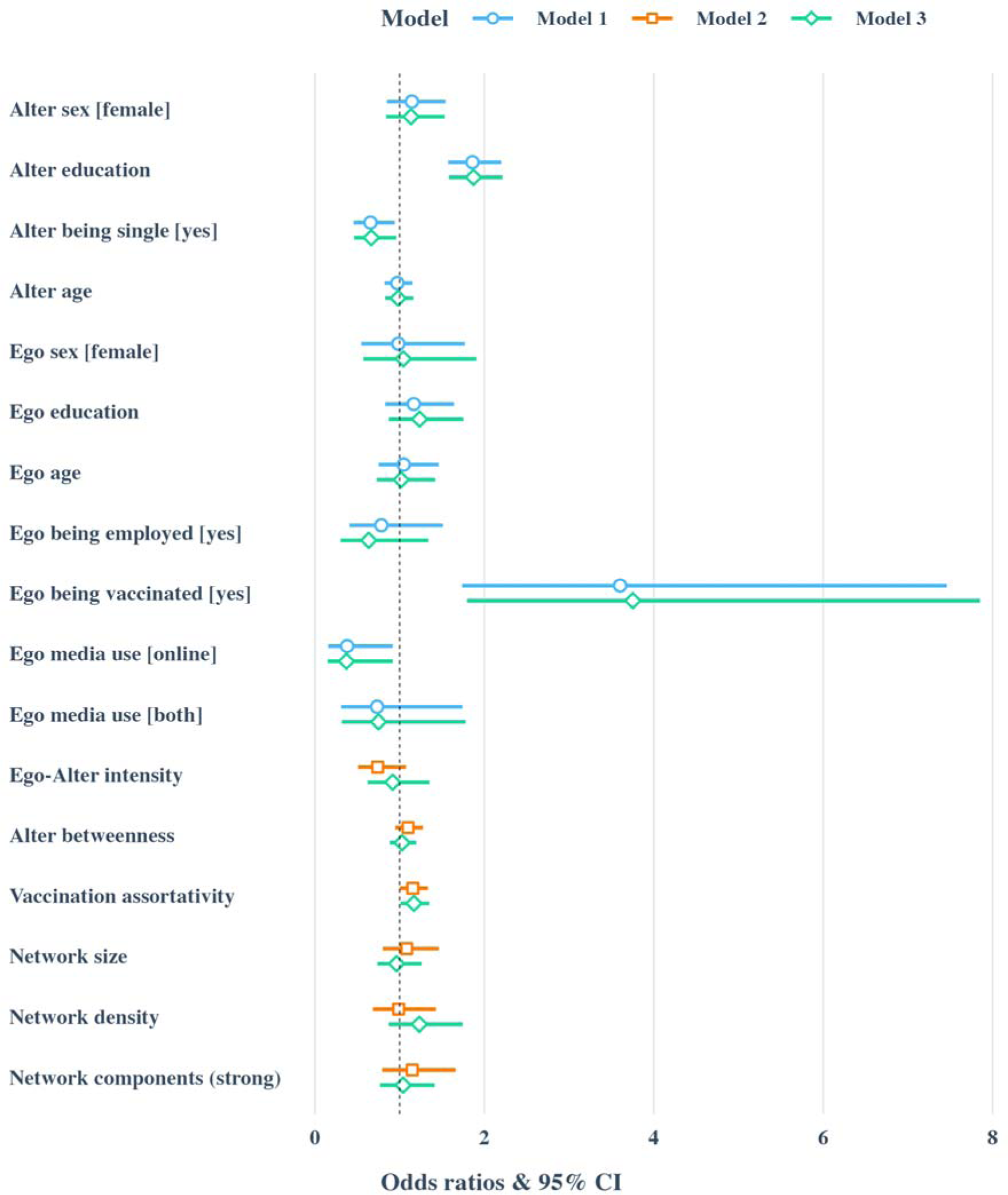
Comparison of mixed multilevel logistic regression models Besides the multilevel logistic regression models, we also performed a logistic general linear model (GLM) with robust standard errors (i.e., standard errors clustered by egos). Table S24 from MA1 presents these results. Comparing the full model from Table 2 with the full model from Table S24 (GLM Model 4), we observe similar results with regards to alters’ education (OR 1.82, 95% CI 1.46-2.26; *P*<.001), egos being vaccinated (OR 1.34, 95% CI 1.06- 1.68; *P*=.013), egos using online media for information about health (OR 0.66, 95% CI 0.44-0.97; *P*=.04), and alters’ vaccination assortativity (OR 1.21, 95% CI 1.01-1.46; *P*=.04). To account for not having random ego intercepts, this model also included the proportion of vaccinated alters minus the alter of interest. For this predictor, also, we observed a positive and significant effect (OR 2.48, 95% CI 2.05-3.01; *P*<.001).

## Discussion

### Principal results

Our study investigated the impact of online media consumption on COVID-19 vaccination behaviors using a PNA approach. We aimed to explore the influence of real-world network structures and compositions on vaccine hesitancy, with a particular focus on the role of individuals’ (egos’) reliance on online media for health and prevention information as the primary predictor. Through the application of mixed multilevel logistic regression models, we assessed the vaccination status of individuals (alters) within 61 personal networks. Our findings indicate that alters within networks of egos who exclusively utilize online media for health and prevention information are less likely to be vaccinated compared to those in networks where egos depend on traditional media sources. Additionally, for alters in networks where egos access both online and traditional media, the effect on vaccination status was not statistically significant. Our results are in line with previous studies that suggest social media use lowers COVID-19 vaccine acceptance [13,19]. The results of the mixed multilevel logistic regression are also supported by general linear models using robust standard errors.

Also, our results support European-level statistics, which indirectly measure digital health literacy. In Romania, in 2023, the proportion of individuals who had at least basic digital skills was 31.09% [53], yet internet usage for social media platforms was at 84.35% [54], and only 32.83% reported seeing in online spaces information which was at least doubtful (compared with the EU average of 53.67%) [55]. Secondary results from our analyses relate to some of the alters’ sociodemographic characteristics and egos’ vaccination status. Alters’ education and being in a relationship increased their odds of being vaccinated, as found in systematic reviews and meta-analyses [66,67]. Similarly, being in the personal network of an ego who is vaccinated increased the odds of alters being vaccinated, which is in line with another PNA study on a sample of Romanian respondents, testing for the assortative mixing of opinions on the COVID-19 vaccine and finding a positive association [31]. In this context, online misinformation and disinformation campaigns can find fertile ground and be enhanced by social influence in real-world networks when they mirror such opinions.

The ways in which individuals use the internet are diverse, and so are the effects of information obtained through this medium when it comes to health opinions and behaviors. For example, Moon et al. [68] found that while a higher frequency of social media use increased the odds of hesitancy, the higher frequency of internet use (for all purposes) decreased them. Allington et al. [18] found that social media usage loses its effect on hesitancy when controlling for trust in government, health professionals, and scientists, and COVID-19 is perceived as a risk. In our models, using both types of media did not differ from using only traditional media, presenting a non-significant effect in predicting alters’ vaccination status. On one hand, this could indicate that exposure to multiple information sources can decrease hesitancy. On the other, this also indicates exogenous factors not taken into account when discussing online behavior. In our supplementary analyses, the media used by egos for health and prevention is not associated with their vaccination status. This could be due to the small sample size of 61 respondents.

An additional finding from our study highlights the impact of assortative mixing on vaccination outcomes. Specifically, alters who had a higher proportion of vaccinated direct network neighbors, relative to the vaccination rate of the entire network, were more likely to be vaccinated themselves. This finding aligns with other research employing PNA to examine assortative mixing concerning attitudes towards COVID-19 vaccination [31] and processed food consumption [62]. Such alignment underlines the potential for further exploratory and comparative studies. Notably, our additional analyses reveal that networks characterized by lower vaccination assortativity tend to belong to unvaccinated egos, particularly those relying solely on online media for information and unvaccinated alters. This pattern suggests a fragmentation in personal networks concerning health beliefs and behaviors, possibly exacerbated by the diverse opinions and information sources about COVID-19 vaccines, echoing the World Health Organization’s concept of an “infodemic” [9]. The fact that the lower assortativity scores are found inside personal networks of unvaccinated egos who use only online media can be further hypothesized as an indicator of reinforcement of online misinformation through network contagion.

Given the limited evidence about the effects of real-world networks on vaccination hesitancy in relation to online media use, it remains an open question whether vaccine hesitancy is the result of assortative mixing (homogeneity) or social mixing (heterogeneity) when we look at networks’ compositions and structures based on nodes’ media use. Building on the available literature about network influences on health behaviors and opinions [31,43,50,51], the working hypothesis would be that assortative mixing on multidimensional aspects of online behavior (e.g., used platforms, followed accounts, eHealth literacy) would positively correlate with assortative mixing on vaccination status. In our results, the similarity between egos and their alters vaccination status, coupled with the fact the elicited alters were not randomly selected by respondents, can be an indicator of this association. The study of personal networks presents the possibility of the *false consensus effect* or *egocentric attribution bias*, wherein individuals attribute to others opinions and behaviors similar to their own [69]. This also can add to our conclusions, as individuals can be influenced in their behavior not by what others do but by what they perceive as being the others’ behaviors and opinions.

The generalization of our findings warrants careful consideration due to the distinct methodological and contextual facets of our study. The data, collected from a rural community in Romania (Eastern Europe) using a link-tracing sampling methodology, provides insights into the dynamics of online media influence on vaccination behaviors within personal networks. However, the rural setting and the specific socio-cultural context of Eastern Europe might limit the direct applicability of our results to urban settings or other regions with different digital literacy levels and media consumption patterns. Nevertheless, it is important to underline that our findings likely reflect the situation in rural areas throughout Eastern Europe, thereby enhancing the inclusivity and relevance of our study within this broader regional context. Future research could benefit from applying similar methodological approaches in diverse settings to explore the universality and variability of these patterns across different social, cultural, and geographical contexts.

The implications raised by our study are not confined to COVID-19 vaccine hesitancy but vaccine hesitancy in general, in offline and online spaces, as real-world personal networks combine both. Further insights on how the information obtained by individuals from online spaces spreads and is reinforced through personal networks are needed, especially for countries low in general literacy, digital literacy, and eHealth literacy. They are more vulnerable in the face of future pandemics disinformation campaigns regarding other vaccines and need communication campaigns and vaccination programs adapted to community-specific patterns.

## Limits

Our study comes with several limitations. Some of them are inherent to the PNA design itself. The information about alters’ attributes and alter-alter relations is reported by the ego, which can lead to inaccuracies [70]. Therefore, the data generation process requires a careful interpretation of the results. To mitigate the impact of inaccuracies in ego-reported data, we limited the number of alters to 25, used communication or meeting frequency as a criterion for the name generator, and the ego-alter relation intensity (emotional closeness and meeting frequency) as a control variable.

Furthermore, we must consider challenges related to our primary variable of interest: the type of media used for health and prevention information. First, this variable was assessed solely at the ego level, because it can lead to potential inaccuracies when egos report on their alters’ media usage. Such inaccuracies could be more pronounced compared to reporting on demographic information (e.g., age, education level, marital status) or vaccination status against COVID-19. Second, our analysis does not differentiate between social media platforms and other online media sources, such as news aggregators (e.g., Google News, Yahoo News). Prior research indicates divergent impacts on vaccine hesitancy and acceptance between these sources, with social media use often correlated with increased vaccine hesitancy, whereas broader internet use or reliance on online news platforms tends to be associated with higher vaccine acceptance. [20,27,68]. The conflation of social media with other online media in our respondents’ reports precludes a nuanced analysis of these distinct influences within our dataset.

Also, the link-trancing sampling method can introduce selection biases regarding respondents and individuals recommended for further participation. The size of the sample, of 61 personal networks with valid data on all variables of interest, can also skew the results. Despite these limitations, the presence of assortative mixing regarding COVID-19 vaccine opinions was also identified in another study with a larger sample size of 443 Romanian respondents, employing a different sampling strategy. This consistency across studies suggests the robustness of our findings despite the methodological challenges [31].

## Conclusions

Our study suggests that health and prevention information disseminated through online media, including social media, significantly impacts individuals’ vaccination status within real-world personal networks. In addition to assortative mixing effects, our findings highlight the positive influence of education on vaccination status. We advocate for a deeper exploration of the composition and structure of personal networks, emphasizing the need to understand individuals’ online behaviors and digital literacy levels for designing effective vaccination communication campaigns and interventions. Romania presents a case study of particular interest, where low digital literacy and vaccination rates coincide with high social media usage, underscoring the challenges of combating online misinformation and disinformation in similar contexts.

## Acknowledgments

The authors kindly thank Dr. Cosmina Cioroboiu, Dr. Bianca Cucos, Florin Găină, Simona- Elena Puncioiu, and Bogdan-Adrian Vidrascu for their valuable contributions to this study.

IO, M-GH, MG., B-EP-M, and IT were supported by the 4P-CAN project, HORIZON-MISS- 2022-CANCER-01, project ID 101104432, programme HORIZON; M-GH was supported by The Research Institute of the University of Bucharest (ICUB) (grant 8405 / 22.07.2023); JL was supported by Deutsche Forschungsgemeinschaft (DFG 321869138); M.P. was supported by the Slovenian Research and Innovation Agency (Javna agencija za znanstvenoraziskovalno in inovacijsko dejavnost Republike Slovenije) (grant P1-0403). The funding sources had no involvement in study design, in the collection, analysis, and interpretation of data, in writing the paper, and in deciding to submit the article for publication.

IO, M-GH, JL, MP, JLM, and MG conceptualized and designed the study. IO, M-GH, B-EP- M, MG, and IT were involved in the investigation process. IO, M-GH, B-EP-M, and IT curated the data. JL designed the analysis framework and formulas. IO, M-GH, JL, MP, JLM, MG and CE drafted the manuscript. IO and M-GH analyzed the data. JL, MP, and JLM validated the analyses.

All authors approved the final version of the manuscript and accepted full responsibility for all aspects of the work described. The authors alone are responsible for the views expressed in this manuscript. In the case of CE, these views do not necessarily represent the decisions, policy or views of the International Agency for Research on Cancer / World Health Organization.

## Abbreviations

COVID-19: coronavirus disease 2019
EU: European Union
GLM: general linear model
ICC: intraclass correlation coefficient
InoMed: Center for Innovation in Medicine
MA1: Multimedia Appendix 1
OR: odds ratio
PNA: personal network analysis
SAGE: Strategic Advisory Group of Experts on Immunization
SARS-CoV-2: severe acute respiratory syndrome coronavirus2
SNA: social network analysis
WHO: World Health Organization

## Data availability

The code and the dataset analyzed in the current study are made openly available in the Zenodo data repository as Oană, I., Hâncean, M.-G., Perc, M., Lerner, J., Pintoiu-Mihăilă, B.- E., Geantă, M., Molina, J.L., Tincă, I., Espina, C. (2024). Replication data for: Online media use and COVID-19 vaccination in real-world personal networks. Zenodo. https://doi.org/10.5281/zenodo.10732217.

## References

1. United Nations. World must be ready to respond to next pandemic: WHO chief. UN News 2023 May 22; Available from: https://news.un.org/en/story/2023/05/1136912 [accessed Jan 15, 2024]

2. Grills LA, Wagner AL. The impact of the COVID-19 pandemic on parental vaccine hesitancy: A cross-sectional survey. Vaccine. 2023 Sep 22;41(41):6127–6133. doi: 10.1016/j.vaccine.2023.08.044

3. Vraga EK, Brady SS, Gansen C, Khan EM, Bennis SL, Nones M, Tang R, Srivastava J, Kulasingam S. A review of HPV and HBV vaccine hesitancy, intention, and uptake in the era of social media and COVID-19. eLife. 2023 Aug 18;12:e85743. doi: 10.7554/eLife.85743

4. The Lancet Child & Adolescent Health. Vaccine hesitancy: a generation at risk. Lancet Child Adolesc Health. 2019 May 1;3(5):281. doi: 10.1016/S2352-4642(19)30092-6

5. MacDonald NE, SAGE Working Group on Vaccine Hesitancy. Vaccine hesitancy: Definition, scope and determinants. Vaccine. 2015 Aug 14;33(34):4161–4164. doi: 10.1016/j.vaccine.2015.04.036

6. Callender D. Vaccine hesitancy: More than a movement. Hum Vaccin Immunother. 2016 Sep;12(9):2464–2468. doi: 10.1080/21645515.2016.1178434

7. Porter D, Porter R. The politics of prevention: Anti-vaccinationism and public health in nineteenth-century England. Med Hist. 1988 Jul;32(3):231–252. doi: 10.1017/S0025727300048225

8. World Health Organization. Munich Security Conference. 2020. Available from: https://www.who.int/director-general/speeches/detail/munich-security-conference [accessed Jan 17, 2024]

9. World Health Organization. Infodemic. Available from: https://www.who.int/health-topics/infodemic#tab=tab_1 [accessed Jan 17, 2024]

10. Puri N, Coomes EA, Haghbayan H, Gunaratne K. Social media and vaccine hesitancy: new updates for the era of COVID-19 and globalized infectious diseases. Hum Vaccin Immunother. 2020 Nov 1;16(11):2586–2593. doi: 10.1080/21645515.2020.1780846

11. Zhao S, Hu S, Zhou X, Song S, Wang Q, Zheng H, Zhang Y, Hou Z. The Prevalence, Features, Influencing Factors, and Solutions for COVID-19 Vaccine Misinformation: Systematic Review. JMIR Public Health Surveill. 2023 Jan 11;9:e40201. doi: 10.2196/40201

12. Rathje S, He JK, Roozenbeek J, Van Bavel JJ, van der Linden S. Social media behavior is associated with vaccine hesitancy. PNAS Nexus. 2022 Sep 30;1(4):pgac207. doi: 10.1093/pnasnexus/pgac207

13. Mascherini M, Nivakoski S. Social media use and vaccine hesitancy in the European Union. Vaccine. 2022 Mar 25;40(14):2215–2225. doi: 10.1016/j.vaccine.2022.02.059

14. Ruiz JB, Bell RA. Predictors of intention to vaccinate against COVID-19: Results of a nationwide survey. Vaccine. 2021 Feb 12;39(7):1080–1086. doi: 10.1016/j.vaccine.2021.01.010

15. Allington D, McAndrew S, Moxham-Hall VL, Duffy B. Media usage predicts intention to be vaccinated against SARS-CoV-2 in the US and the UK. Vaccine. 2021 Apr 28;39(18):2595–2603. doi: 10.1016/j.vaccine.2021.02.054

16. Tollison AC, LoPresti A. Media consumption and COVID-19 vaccination hesitancy: health literacy as a response. Commun Res Rep. 2023 Oct 19;40(5):272–282. doi: 10.1080/08824096.2023.2270904

17. Carrieri V, Guthmuller S, Wübker A. Trust and COVID-19 vaccine hesitancy. Sci Rep. 2023 Jun 7;13(1):9245. doi: 10.1038/s41598-023-35974-z

18. Allington D, McAndrew S, Moxham-Hall V, Duffy B. Coronavirus conspiracy suspicions, general vaccine attitudes, trust and coronavirus information source as predictors of vaccine hesitancy among UK residents during the COVID-19 pandemic. Psychol Med. 2023 Jan;53(1):236–247. doi: 10.1017/S0033291721001434

19. Jennings W, Valgarðsson V, McKay L, Stoker G, Mello E, Baniamin HM. Trust and vaccine hesitancy during the COVID-19 pandemic: A cross-national analysis. Vaccine X. 2023 Aug;14:100299. doi: 10.1016/j.jvacx.2023.100299

20. Viswanath K, Bekalu M, Dhawan D, Pinnamaneni R, Lang J, McLoud R. Individual and social determinants of COVID-19 vaccine uptake. BMC Public Health. 2021 Apr 28;21(1):818. doi: 10.1186/s12889-021-10862-1

21. Ouyang H, Ma X, Wu X. The prevalence and determinants of COVID-19 vaccine hesitancy in the age of infodemic. Hum Vaccin Immunother. 2022 Dec 31;18(1):2013694. doi: 10.1080/21645515.2021.2013694

22. Cascini F, Pantovic A, Al-Ajlouni YA, Failla G, Puleo V, Melnyk A, Lontano A, Ricciardi W. Social media and attitudes towards a COVID-19 vaccination: A systematic review of the literature. EClinicalMedicine. 2022 Jun;48:101454. doi: 10.1016/j.eclinm.2022.101454

23. McKinley CJ, Limbu Y. Promoter or barrier? Assessing how social media predicts Covid-19 vaccine acceptance and hesitancy: A systematic review of primary series and booster vaccine investigations. Soc Sci Med. 2024 Jan;340:116378. doi: 10.1016/j.socscimed.2023.116378

24. Kafadar AH, Tekeli GG, Jones KA, Stephan B, Dening T. Determinants for COVID-19 vaccine hesitancy in the general population: a systematic review of reviews. J of Public Health. 2023 Nov 1;31:1829–1845. doi: 10.1007/s10389-022-01753-9

25. Gewirtz-Meydan A, Mitchell K, Shlomo Y, Heller O, Grinstein-Weiss M. COVID-19 Among Youth in Israel: Correlates of Decisions to Vaccinate and Reasons for Refusal. J Adolesc Health. 2022 Mar;70(3):396–402. doi: 10.1016/j.jadohealth.2021.11.016

26. Luo C, Chen A, Cui B, Liao W. Exploring public perceptions of the COVID-19 vaccine online from a cultural perspective: Semantic network analysis of two social media platforms in the United States and China. Telemat Inform. 2021 Dec;65:101712. doi: 10.1016/j.tele.2021.101712

27. Liu PL, Zhao X, Wan B. COVID-19 information exposure and vaccine hesitancy: The influence of trust in government and vaccine confidence. Psychol Health Med. 2023 Jan;28(1):27–36. doi: 10.1080/13548506.2021.2014910

28. Tan M, Straughan PT, Cheong G. Information trust and COVID-19 vaccine hesitancy amongst middle-aged and older adults in Singapore: A latent class analysis Approach. Soc Sci Med. 2022 Mar;296:114767. doi: 10.1016/j.socscimed.2022.114767

29. Lee M, You M. Direct and Indirect Associations of Media Use With COVID-19 Vaccine Hesitancy in South Korea: Cross-sectional Web-Based Survey. J Med Internet Res. 2022 Jan 6;24(1):e32329. doi: 10.2196/32329

30. Klaus C, Wascher M, KhudaBukhsh WR, Tien JH, Rempała GA, Kenah E. Assortative mixing among vaccination groups and biased estimation of reproduction numbers. Lancet Infect Dis. 2022 May;22(5):579–581. doi: 10.1016/S1473-3099(22)00155-4

31. Hâncean M-G, Lerner J, Perc M, Molina JL, Geantă M. Assortative mixing of opinions about COVID-19 vaccination in personal networks. Sci Rep. 2024 Feb 9;14(1):3385. doi: 10.1038/s41598-024-53825-3

32. McCarty C, Lubbers MJ, Vacca R, Molina JL. Conducting personal network research: a practical guide. New York City: The Guilford Press; 2019. ISBN:978-1-4625-3838-6

33. Hwang J, Su M-H, Jiang X, Lian R, Tveleneva A, Shah D. Vaccine discourse during the onset of the COVID-19 pandemic: Topical structure and source patterns informing efforts to combat vaccine hesitancy. PLoS One. 2022 Jul 27;17(7):e0271394. doi: 10.1371/journal.pone.0271394

34. Bonifazi G, Breve B, Cirillo S, Corradini E, Virgili L. Investigating the COVID-19 vaccine discussions on Twitter through a multilayer network-based approach. Inf Process Manag. 2022 Nov;59(6):103095. doi: 10.1016/j.ipm.2022.103095

35. Muric G, Wu Y, Ferrara E. COVID-19 Vaccine Hesitancy on Social Media: Building a Public Twitter Data Set of Antivaccine Content, Vaccine Misinformation, and Conspiracies. JMIR Public Health Surveill. 2021 Nov 17;7(11):e30642. doi: 10.2196/30642

36. Wang D, Zhou Y, Ma F. Opinion Leaders and Structural Hole Spanners Influencing Echo Chambers in Discussions About COVID-19 Vaccines on Social Media in China: Network Analysis. J Med Internet Res. 2022 Nov 18;24(11):e40701. doi: 10.2196/40701

37. Li Y, Gee W, Jin K, Bond R. Examining Homophily, Language Coordination, and Analytical Thinking in Web-Based Conversations About Vaccines on Reddit: Study Using Deep Neural Network Language Models and Computer-Assisted Conversational Analyses. J Med Internet Res. 2023 Mar 23;25:e41882. doi: 10.2196/41882

38. Hâncean M-G, Lerner J, Perc M, Ghită MC, Bunaciu D-A, Stoica AA, Mihăilă B-E. The role of age in the spreading of COVID-19 across a social network in Bucharest. J Complex Netw. 2021 Sep 7;9(4). doi: 10.1093/comnet/cnab026

39. Hâncean M-G, Lerner J, Perc M, Oană I, Bunaciu D-A, Stoica AA, Ghită M-C. Occupations and their impact on the spreading of COVID-19 in urban communities. Sci Rep. 2022 Aug 18;12(1):14115. doi: 10.1038/s41598-022-18392-5

40. Hâncean M-G, Ghită MC, Perc M, Lerner J, Oană I, Mihăilă B-E, Stoica AA, Bunaciu D-A. Disaggregated data on age and sex for the first 250 days of the COVID-19 pandemic in Bucharest, Romania. Sci Data. 2022 May 31;9(1). doi: 10.1038/s41597-022-01374-7

41. Cohen-Cole E, Fletcher JM. Is obesity contagious? Social networks vs. environmental factors in the obesity epidemic. J Health Econ. 2008 Sep;27(5):1382–1387. doi: 10.1016/j.jhealeco.2008.04.005

42. Aral S, Muchnik L, Sundararajan A. Distinguishing influence-based contagion from homophily-driven diffusion in dynamic networks. Proc Natl Acad Sci U S A. 2009 Dec 22;106(51):21544–21549. doi: 10.1073/pnas.0908800106

43. Christakis NA, Fowler JH. Social contagion theory: examining dynamic social networks and human behavior. Stat Med. 2013 Feb 20;32(4):556–577. doi: 10.1002/sim.5408

44. McPherson M, Smith-Lovin L, Cook JM. Birds of a Feather: Homophily in Social Networks. Annu Rev Sociol. 2001 Aug 1;27(1):415–444. doi: 10.1146/annurev.soc.27.1.415

45. Hill AL, Rand DG, Nowak MA, Christakis NA. Infectious Disease Modeling of Social Contagion in Networks. PLoS Comput Biol. 2010 Nov 4;6(11):e1000968. doi: 10.1371/journal.pcbi.1000968

46. Centola D. An experimental study of homophily in the adoption of health behavior. Science. 2011 Dec 2;334(6060):1269–1272. doi: 10.1126/science.1207055

47. Kumar P, McCafferty L, Dhand A, Rao S, Díaz-Valdés A, Tabak RG, Brownson RC, Yadama GN. Association of personal network attributes with clean cooking adoption in rural South India. Environ Res Lett. 2021 Jun 16;16(6):064087. doi: 10.1088/1748-9326/ac0746

48. Pollack CE, Soulos PR, Herrin J, Xu X, Christakis NA, Forman HP, Yu JB, Killelea BK, Wang S-Y, Gross CP. The Impact of Social Contagion on Physician Adoption of Advanced Imaging Tests in Breast Cancer. J Natl Cancer Inst. 2017 Aug 1;109(8):djw330. doi: 10.1093/jnci/djw330

49. Yu JB, Pollack CE, Herrin J, Soulos PR, Zhu W, Xu X, Gross CP. Peer Influence on Physician Use of Shorter Course External Beam Radiation Therapy for Patients with Breast Cancer. Pract Radiat Oncol. 2020 Apr;10(2):75–83. doi: 10.1016/j.prro.2019.11.001

50. Barclay VC, Smieszek T, He J, Cao G, Rainey JJ, Gao H, Uzicanin A, Salathé M. Positive Network Assortativity of Influenza Vaccination at a High School: Implications for Outbreak Risk and Herd Immunity. PLoS One. 2014 Feb 5;9(2):e87042. doi: 10.1371/journal.pone.0087042

51. Ni L, Chen Y, de Brujin O. Towards understanding socially influenced vaccination decision making: An integrated model of multiple criteria belief modelling and social network analysis. Eur J Oper Res. 2021 Aug 16;293(1):276–289. doi: 10.1016/j.ejor.2020.12.011

52. Mathieu E, Ritchie H, Ortiz-Ospina E, Roser M, Hasell J, Appel C, Giattino C, Rodés-Guirao L. A global database of COVID-19 vaccinations. Nat Hum Behav. 2021 Jul;5(7):947–953. doi: 10.1038/s41562-021-01122-8

53. Eurostat. Individuals’ level of digital skills (from 2021 onwards). 2023. Available from: https://ec.europa.eu/eurostat/databrowser/view/isoc_sk_dskl_i21/default/table?lang=en [accessed Jan 15, 2024]

54. Eurostat. Individuals - internet activities. 2023. Available from: https://ec.europa.eu/eurostat/databrowser/view/isoc_ci_ac_i/default/table?lang=en [accessed Jan 15, 2024]

55. Eurostat. Evaluating data, information and digital content (2021 onwards). 2023. Available from: https://ec.europa.eu/eurostat/databrowser/view/isoc_sk_edic_i21/default/table?lang=en [accessed Jan 15, 2024]

56. 4P-CAN. Living Labs. 2024. Available from: https://4p-can.eu/living-labs/#romania [accessed Jan 17, 2024]

57. Hâncean M-G, Lubbers MJ, Molina JL. Measuring transnational social fields through binational link-tracing sampling. PLoS One. 2021 Jun 14;16(6):e0253042. doi: 10.1371/journal.pone.0253042

58. Molina JL, Lubbers MJ, Hâncean M-G, Fradejas-García I. Short Take: Sampling from Transnational Social Fields. Field Methods. 2022 Aug 1;34(3):256–264. doi: 10.1177/1525822X221105920

59. Complex Data Collective. Network Canvas: Interviewer. Zenodo; 2023. doi: 10.5281/ZENODO.6026548

60. Scott J. Social Network Analysis: A Handbook. London: SAGE Publications Ltd; 2000. ISBN:978-0-7619-6339-4

61. Wasserman S, Faust K. Social Network Analysis: Methods and Applications. Cambridge: Cambridge University Press; 1994. ISBN:0-521-38707-8

62. Hâncean M-G, Lerner J, Perc M, Molina JL, Geantă M, Oană I, Pintoiu-Mihăilă B-E, Puncioiu S-E. Processed food intake assortativity in the personal networks of East European older adults. medRxiv. Preprint posted online January 26, 2024. doi: 10.1101/2024.01.25.24301787

63. Borgatti SP, Everett MG, Johnson JC. Analyzing Social Networks. London: SAGE Publications Ltd; 2013. ISBN:978-1-4462-4740-2

64. Gelman A, Hill J. Data analysis using regression and multilevel/hierarchical models. Cambridge: Cambridge University Press; 2007. ISBN:978-0-521-86706-1

65. Vacca R. Multilevel models for personal networks: methods and applications. Ital J Appl Stat. 2018 Apr;30(1):59–97. doi: 10.26398/IJAS.0030-003

66. Adu P, Popoola T, Medvedev ON, Collings S, Mbinta J, Aspin C, Simpson CR. Implications for COVID-19 vaccine uptake: A systematic review. J Infect Public Health. 2023 Mar;16(3):441–466. doi: 10.1016/j.jiph.2023.01.020

67. Baghani M, Fathalizade F, Loghman AH, Samieefar N, Ghobadinezhad F, Rashedi R, Baghsheikhi H, Sodeifian F, Rahimzadegan M, Akhlaghdoust M. COVID-19 vaccine hesitancy worldwide and its associated factors: a systematic review and meta-analysis. Sci One Health. 2023 Jan 1;2:100048. doi: 10.1016/j.soh.2023.100048

68. Moon I, Han J, Kim K. Determinants of COVID-19 vaccine Hesitancy: 2020 California Health Interview Survey. Prev Med Rep. 2023 Jun;33:102200. doi: 10.1016/j.pmedr.2023.102200

69. Ross L, Greene D, House P. The “false consensus effect”: An egocentric bias in social perception and attribution processes. J Exp Soc Psychol. 1977 May;13(3):279–301. doi: 10.1016/0022-1031(77)90049-X

70. Wellman B. Challenges in Collecting Personal Network Data: The Nature of Personal Network Analysis. Field Methods. 2007 May 1;19(2):111–115. doi: 10.1177/1525822X06299133

